# Effectiveness of influenza vaccination against infection in UK healthcare workers during winter 2023-24: the SIREN cohort study

**DOI:** 10.1101/2025.06.07.25329188

**Authors:** Luke J McGeoch, Sarah Foulkes, Heather Whitaker, Katie Munro, Jameel Khawam, Dominic Sparkes, Andre Charlett, Colin S Brown, Ana Atti, Jasmin Islam, Susan Hopkins, Nick Andrews, Victoria J Hall

## Abstract

**Objectives:** To determine vaccine effectiveness against influenza infection among UK healthcare workers between 1 September 2023 and 31 March 2024.

**Methods:** We conducted a prospective cohort study, including hospital-based healthcare workers (HCWs) enrolled in the SARS-CoV-2 Immunity & Reinfection Evaluation (SIREN) study. Participants completed fortnightly influenza PCR testing and questionnaires. Influenza vaccination status was identified from national vaccination records and questionnaires. Vaccine effectiveness against PCR-positive influenza was estimated using Cox regression adjusted for age group, sex, chronic disease status, patient-facing role, and region. Case-control and test-negative case-control (TNCC) analyses, using multivariable logistic regression, were also performed.

**Results** Of 4,934 participants, most were female (78.7%), aged 35-64 years (88.6%), and white (85.6%). Overall, 3,857 (78.2%) received influenza vaccination and 266 (5.4%) tested positive for influenza, of which 227 (85.3%) reported acute respiratory infection symptoms. Adjusted vaccine effectiveness was 39.9% (95% confidence interval 21.8 – 53.8), and similar using case-control (41.2%, 22.5 – 55.2) and TNCC (45.9%, 21.8 – 62.2) approaches.

**Conclusions:** Influenza vaccine effectiveness was 40%, consistent with estimates for symptomatic patients. Applied to the combined UK healthcare workforce, this potentially translates to prevention of over 50,000 infections. These findings emphasise the importance of seasonal influenza vaccination to reduce healthcare workers infections and thereby protect patients and reduce workforce pressures.

**HIGHLIGHTS:**

- A large cohort of over 4,900 healthcare workers (HCWs) enrolled in the SIREN study completed fortnightly influenza PCR testing and questionnaires throughout the winter 2023-24 season.
- One in twenty HCWs were infected with influenza, including 4.6% of vaccinated HCWs and 8.1% of unvaccinated HCWs, and 85% experienced acute respiratory infection symptoms.
- Healthcare workers who received influenza vaccination were 40% less likely to become infected with influenza compared to those who were not vaccinated.

## INTRODUCTION

In the United Kingdom, like many countries, annual seasonal influenza vaccination is routinely offered to groups considered to be at higher risk of influenza infection and adverse disease outcomes [1]. Healthcare workers (HCWs) with patient contact are included in this group to reduce their personal infection risk, transmission to patients and workforce absence [2, 3]. For the 2023-24 season, HCWs were eligible for vaccination from 1 September 2023, with the majority receiving either the cell-based quadrivalent influenza vaccine (QIVc) or recombinant quadrivalent influenza vaccine (QIVr) [2]. Though an aim has been set for 75-80% vaccination coverage among HCWs with patient contact in England [4], uptake declined from a peak of 76.8% in 2020-21 to 42.8% in 2023-24 [3]. The same trend has been observed in Scotland, Wales and Northern Ireland [5–7]. Perceptions of limited personal infection risk, poor vaccine effectiveness and adverse vaccine effects contribute to low uptake [8].

A small number of randomised controlled trials and cohort studies, each restricted to a single or small number of sites, have investigated the effectiveness of influenza vaccination in HCWs [9–11]. Ongoing surveillance of influenza vaccine effectiveness (VE) predominantly relies on test-negative case-control (TNCC) studies including symptomatic members of the general population attending healthcare settings and receiving one-off testing [12]. Few studies have performed head-to-head comparisons of different analytical approaches for assessing influenza VE, primarily using simulation-based approaches [13–15].

We aimed to determine VE against all influenza infection among HCWs in the SIREN cohort study who completed fortnightly influenza PCR tests during the 2023-24 influenza season, and to compare VE estimates obtained using cohort, traditional case-control and TNCC analytical approaches.

## METHODS

### Study design

The SARS-CoV-2 Immunity & Reinfection Evaluation (SIREN) study, run by the UK Health Security Agency (UKHSA), is a prospective cohort study of hospital-based National Health Service (NHS) HCWs followed continuously since June 2020, as described previously [16].

### Data sources

At enrolment, participants completed an online study questionnaire obtaining socio-demographic data and chronic disease history. Index of multiple deprivation (IMD), a measure of neighbourhood relative deprivation calculated by the Office of National Statistics, was obtained through linkage with participant postcodes [17].

Between 1 September 2023 and 31 March 2024, enrolled participants completed fortnightly respiratory swabs and online follow-up questionnaires. Questionnaires captured data on symptoms, testing, and exposures (household, community, and occupational). Self-completed nose and oropharyngeal swab kits were posted to participants and returned to a central UKHSA laboratory for reverse transcriptase polymerase–chain reaction (PCR) testing for influenza, SARS-CoV-2, and respiratory syncytial virus (RSV). PCR results were reported to the study team by the central laboratory.

Data on seasonal influenza vaccination during the follow-up period were obtained from follow-up questionnaires and from linkage on personal identifiable information (NHS number, surname, date of birth, and postcode) to national vaccination registries.

### Study population

Individuals enrolled in the SIREN cohort were eligible for this study if they had at least two valid influenza PCR results during the study period. Individuals were excluded if influenza vaccination status was unknown, or if they had a positive influenza test result recorded between 1 April 2023 and 31 August 2023.

### Study variables

The primary outcome was influenza infection, measured by PCR, regardless of symptom status. A secondary outcome was symptomatic influenza infection, defined as an acute respiratory infection (ARI) with onset during the period 7 days either side of the sample date of a positive influenza PCR test, where an ARI episode was defined as a self-report of one or more of cough, sore throat, shortness of breath, or coryza.

Participants were categorised by 2023-24 influenza vaccination status. Given that individuals are not considered to have developed an immunological response to influenza vaccination during the window 0-13 days post-vaccination, vaccination status was assigned as unvaccinated, partially vaccinated (0-13 days) or vaccinated (≥14 days).

Other study variables obtained from questionnaires included age group, sex, ethnic group, IMD, region of residence, presence of any chronic disease or immunosuppression, occupational group, occupational setting, whether the HCW had a patient-facing role, and presence of others in the household.

### Follow-up period

Individual participants were followed up from the sample date of their first respiratory swab on or after 1 September 2023 to either: the sample date of the last negative respiratory swab or, if they tested positive for influenza on PCR during the follow-up period, the sample date of the first positive respiratory swab, following which they were censored. Vaccinated participants contributed to unvaccinated follow-up time prior to the date of vaccination, and to vaccinated follow-up time from 14 days post-vaccination.

### Statistical analysis

The baseline characteristics of participants were described using frequencies and percentages. The main approach used to assess VE was a cohort study, with VE (%) estimated as (1-hazard ratio) x 100 using Cox proportional hazards regression. The proportional hazards assumption was assessed using the Schoenfeld test of the independence between scaled Schoenfeld residuals and time. Covariates included in a multivariable regression model a priori as recognised confounders included: age group, sex, presence of any chronic disease or immunosuppression, patient-facing role, and region of residence. The inclusion of interactions terms between covariates in the model was evaluated using Akaike information criterion values and likelihood-ratio tests to assess goodness of fit. For the analysis of the secondary outcome of symptomatic influenza infection, participants were excluded if they did not have a follow-up questionnaire including symptom information completed within 7 days either side of a respiratory swab sample date.

Additional analytical approaches were used to assess the consistency of VE estimates. A case-control analysis categorised participants as cases if they tested positive for any influenza virus at least once during the follow-up period and as controls if all PCR tests were negative for influenza. A test-negative case-control (TNCC) analysis included participants reporting at least one ARI episode during the study period (since TNCC studies ordinarily include symptomatic patients seeking healthcare services), with restriction to PCR test results that were accompanied by an ARI episode within 7 days either side of the sample date. Using both approaches, for cases, the first positive PCR test result was included in the analysis. For controls, one PCR test result was randomly selected for inclusion in the analysis. VE (%) was estimated as (1-odds ratio) x 100 using logistic regression. Logistic regression models included the same covariates as the cohort analysis, as well as a spline for number of days since follow-up commenced on 1 September 2023, with 4 degrees of freedom.

In each analysis, waning of VE over time was assessed by comparing two distinct post-immunisation follow-up periods (14-104 days, ≥105 days), using a cut-off of 90 days after the post-vaccination window for development of immune protection.

All analyses were conducted using R version 4.4.1 [18].

### Ethics

The SIREN study protocol was approved by the Berkshire Research Ethics Committee on 22/05/2020 [16]. Amendment 31, which covered the current study, received approval on 22/08/2023. The study collected consent from all participants. ISRCTN Registry number: ISRCTN11041050.

### Patient and public involvement

The design, implementation and evaluation of the SIREN study is informed by a Participant involvement Panel (PIP) established in collaboration with the British Society for Immunology and drawn from a range of demographic and professional backgrounds [19]. The PIP provides regular feedback on the SIREN study and is also involved in the co-development of participant and public engagement activities that support the ongoing running of the study.

## RESULTS

### Participants

Overall, 4,934 participants were included in the study (Fig. 1), representing 82.6% of the SIREN cohort under follow-up at the start of the study period. Median age was 55 years (IQR 47-60 years). The majority of participants were female (78.7%) and of white ethnicity (85.6%) (Table 1). Chronic disease or immunosuppression was reported by 26.9% of participants. The most common occupational groups were nurses and healthcare assistants (36.5%), administrative/executive staff (15.9%), and doctors (14.1%). The median follow-up duration per participant was 156 days (IQR 127-178 days). The combined follow-up duration was 2,010 person-years. Participants completed a median of 12 PCR tests (IQR 9-13) and 11 questionnaires (IQR 9-12). The median time between both successive PCR tests and successive questionnaires was 14 days (IQR 13-16), which was consistent between vaccinated and unvaccinated participants. Collectively, participants contributed 49,591 questionnaires and 52,333 PCR tests.

**Fig. 1.**
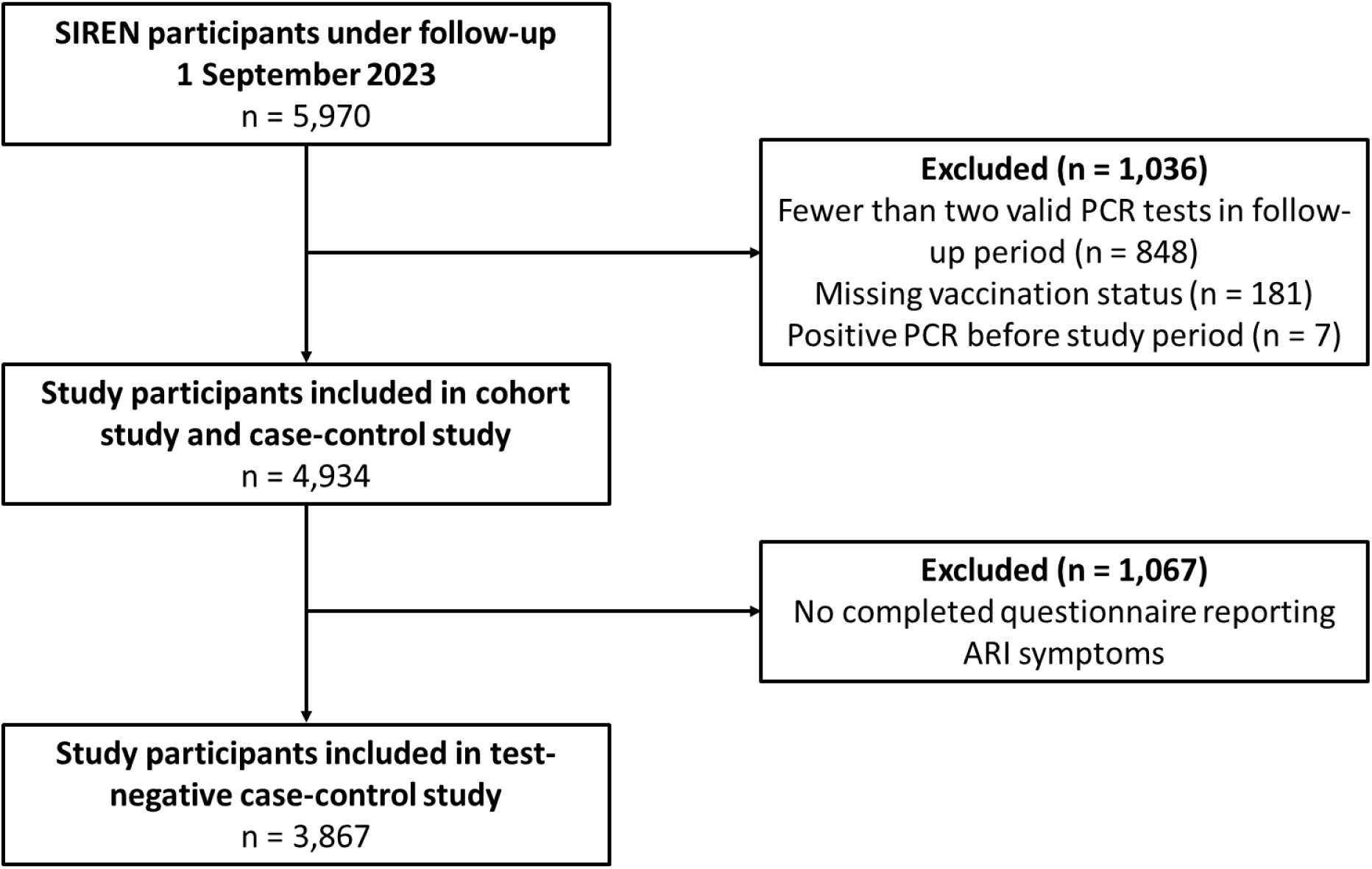
SIREN participants included in the study.

**Table 1.** Baseline characteristics of study participants by 2023-24 seasonal influenza vaccination status. Abbreviations: aQIV, adjuvanted quadrivalent inactivated influenza vaccine; IMD, index of multiple deprivation, QIVc, quadrivalent cell cultured inactivated influenza vaccine; QIVe, quadrivalent inactivated egg-based influenza vaccine; QIVr, recombinant quadrivalent inactivated influenza vaccine.

|  | Vaccinated<br>(n = 3,857) |  | Unvaccinated<br>(n=1,077) |  | Total<br>(n = 4,934) |  |
| --- | --- | --- | --- | --- | --- | --- |
|  | n | % | n | % | n | % |
| **Age group in years** |  |  |  |  |  |  |
| <34 | 134 | 3.5 | 52 | 4.8 | 186 | 3.8 |
| 35-64 | 3,387 | 87.8 | 984 | 91.4 | 4,371 | 88.6 |
| ≥65 | 336 | 8.7 | 41 | 3.8 | 377 | 7.6 |
| **Sex** |  |  |  |  |  |  |
| Male | 870 | 22.6 | 182 | 16.9 | 1,052 | 21.3 |
| Female | 2,987 | 77.4 | 895 | 83.1 | 3,882 | 78.7 |
| **Ethnic group** |  |  |  |  |  |  |
| Black | 98 | 2.5 | 60 | 5.6 | 158 | 3.2 |
| Asian | 259 | 6.7 | 104 | 9.7 | 363 | 7.4 |
| Mixed | 70 | 1.8 | 35 | 3.2 | 105 | 2.1 |
| White | 3,372 | 87.4 | 852 | 79.1 | 4,224 | 85.6 |
| Other | 52 | 1.3 | 22 | 2.0 | 74 | 1.5 |
| Unknown | 6 | 0.2 | 4 | 0.4 | 10 | 0.2 |
| **IMD quintile** |  |  |  |  |  |  |
| 1 (most deprived) | 338 | 8.8 | 116 | 10.8 | 454 | 9.2 |
| 2 | 588 | 15.2 | 231 | 21.4 | 819 | 16.6 |
| 3 | 818 | 21.2 | 230 | 21.4 | 1,048 | 21.2 |
| 4 | 917 | 23.8 | 265 | 24.6 | 1,182 | 24.0 |
| 5 (least deprived) | 1,150 | 29.8 | 228 | 21.2 | 1,378 | 27.9 |
| Unknown | 46 | 1.2 | 7 | 0.6 | 53 | 1.1 |
| **Region** |  |  |  |  |  |  |
| South | 1,023 | 26.5 | 256 | 23.8 | 1,279 | 25.9 |
| London | 418 | 10.8 | 168 | 15.6 | 586 | 11.9 |
| Midlands | 961 | 24.9 | 278 | 25.8 | 1,239 | 25.1 |
| North | 822 | 21.3 | 168 | 15.6 | 990 | 20.1 |
| Scotland | 531 | 13.8 | 172 | 16.0 | 703 | 14.2 |
| Wales | 102 | 2.6 | 35 | 3.2 | 137 | 2.8 |
| **Chronic disease or immunosuppression** |  |  |  |  |  |  |
| No | 2751 | 71.3 | 854 | 79.3 | 3605 | 73.1 |
| Yes | 1,106 | 28.7 | 223 | 20.7 | 1,329 | 26.9 |
| **Occupational group** |  |  |  |  |  |  |
| Administrative or executive, office-based occupation | 631 | 16.4 | 155 | 14.4 | 786 | 15.9 |
| Doctor | 611 | 15.8 | 85 | 7.9 | 696 | 14.1 |
| Health-care scientist | 255 | 6.6 | 69 | 6.4 | 324 | 6.6 |
| Midwife | 118 | 3.1 | 57 | 5.3 | 175 | 3.5 |
| Nonclinical support staff: maintenance staff, security guard, or hospital porter | 71 | 1.8 | 23 | 2.1 | 94 | 1.9 |
| Nursing or health-care assistant | 1,325 | 34.4 | 474 | 44.0 | 1,799 | 36.5 |
| Pharmacist | 97 | 2.5 | 19 | 1.8 | 116 | 2.4 |
| Physiotherapist, occupational therapist, or speech and language therapist | 180 | 4.7 | 52 | 4.8 | 232 | 4.7 |
| Student | 120 | 3.1 | 26 | 2.4 | 146 | 3.0 |
| Other | 449 | 11.6 | 117 | 10.9 | 566 | 11.5 |
| **Occupational setting** |  |  |  |  |  |  |
| Office | 881 | 22.8 | 204 | 18.9 | 1,085 | 22.0 |
| Patient-facing (non-clinical) | 185 | 4.8 | 45 | 4.2 | 230 | 4.7 |
| Outpatient | 776 | 20.1 | 238 | 22.1 | 1,014 | 20.6 |
| Maternity / Labour ward | 61 | 1.6 | 18 | 1.7 | 79 | 1.6 |
| Ambulance / Emergency department | 87 | 2.3 | 23 | 2.1 | 110 | 2.2 |
| Inpatient wards | 462 | 12.0 | 137 | 12.7 | 599 | 12.1 |
| Intensive care | 163 | 4.2 | 46 | 4.3 | 209 | 4.2 |
| Theatres | 99 | 2.6 | 32 | 3.0 | 131 | 2.7 |
| Other | 1,143 | 29.6 | 334 | 31 | 1,477 | 29.9 |
| **Patient contact** |  |  |  |  |  |  |
| No | 674 | 17.5 | 137 | 12.7 | 811 | 16.4 |
| Yes | 3,183 | 82.5 | 940 | 87.3 | 4,123 | 83.6 |
| **Household** |  |  |  |  |  |  |
| Lives alone | 509 | 13.2 | 127 | 11.8 | 636 | 12.9 |
| Lives with other adults | 1,958 | 50.8 | 508 | 47.2 | 2,466 | 50.0 |
| Lives with others, including children | 1,380 | 35.8 | 437 | 40.6 | 1,817 | 36.8 |
| Unknown | 10 | 0.3 | 5 | 0.5 | 15 | 0.3 |
| **Vaccine type** |  |  |  |  |  |  |
| aQIV | 179 | 4.6 | - | - | 179 | 3.6 |
| QIVc | 2881 | 74.7 | - | - | 2881 | 58.4 |
| QIVe | 75 | 1.9 | - | - | 75 | 1.5 |
| QIVr | 198 | 5.1 | - | - | 198 | 4.0 |
| Unknown | 524 | 13.6 | - | - | - | - |
| **Influenza vaccine in 2022-23** |  |  |  |  |  |  |
| No | 223 | 5.8 | 643 | 59.7 | 866 | 17.6 |
| Yes | 3,634 | 94.2 | 434 | 40.3 | 4,068 | 82.4 |
| **Influenza vaccine at least once in 2020-23** |  |  |  |  |  |  |
| No | 66 | 1.7 | 447 | 41.5 | 513 | 10.4 |
| Yes | 3,791 | 98.3 | 630 | 58.5 | 4,421 | 89.6 |

### Vaccination coverage

Overall, 3,857 (78.2%) participants received influenza vaccination. Vaccinations peaked in week 40 of 2023 and 3,745 (97.1%) vaccinations took place prior to 01 December 2023 (Fig. 2A). Vaccine type was known for 86.4% of vaccinated participants, with QIVc being most commonly reported (74.7%). Compared to unvaccinated participants, those vaccinated were more likely to be aged ≥65 years (8.7% versus 3.8%), male (22.6% versus 16.9%), of white ethnicity (87.4% versus 79.1%), and have a chronic disease or immunosuppression (28.7% versus 20.7%). Vaccinated participants were more likely to have received influenza vaccination in the previous influenza season (94.2% versus 40.3%) and at least once during the three preceding influenza seasons (98.3% versus 58.5%).

**Fig. 2.**
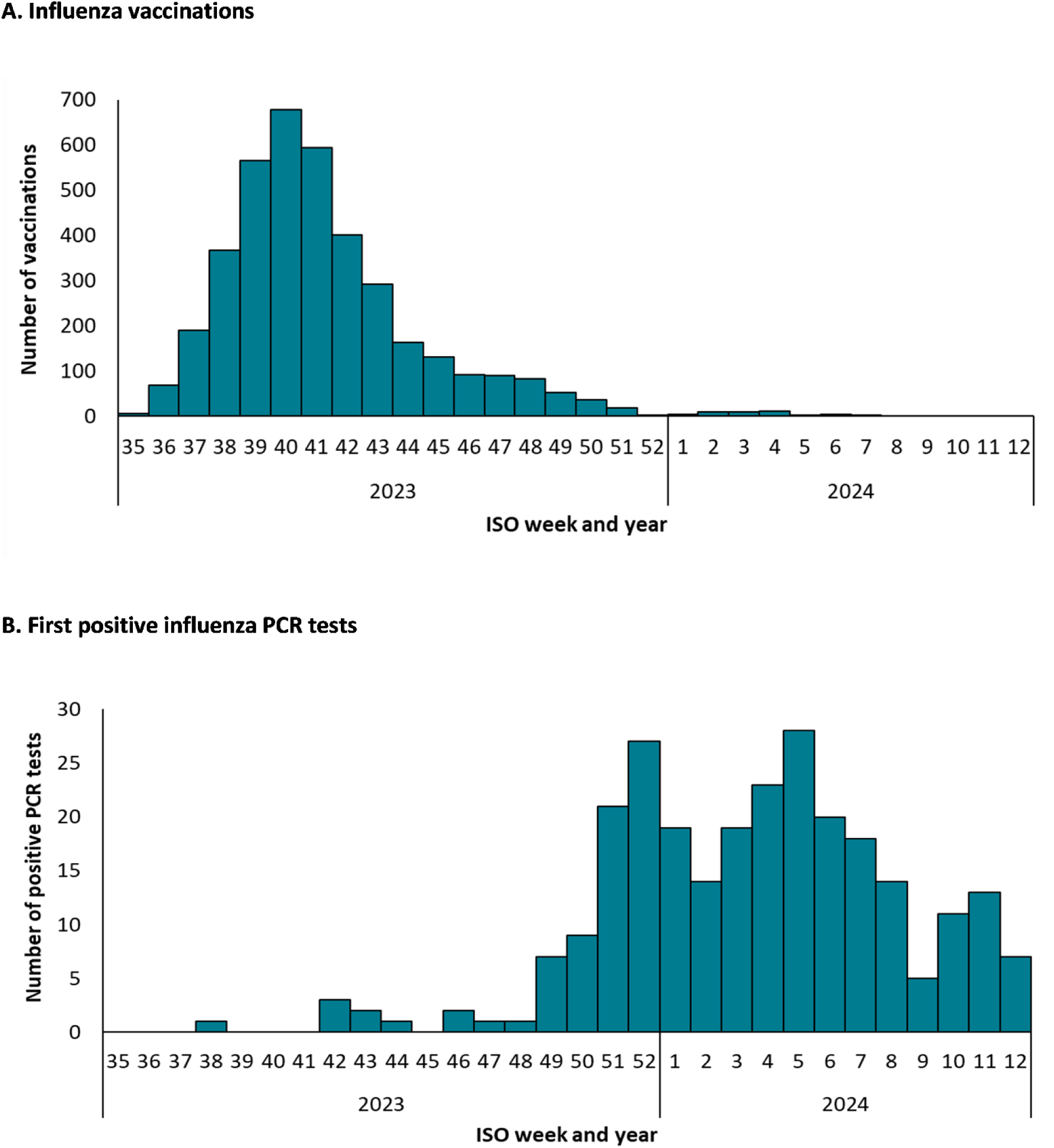
Number of influenza vaccinations (A) and first positive influenza PCR tests (B) by ISO week and year among study participants during the 2023-2024 influenza season.

### Influenza infections

Overall, 266 (5.4%) participants tested positive for influenza, of which 255 (95.9%) had a first positive test result on or after 01 December 2023, and 227 (85.3%) reported an ARI episode within 7 days either side of the respiratory swab sample date. The number of first positive influenza tests peaked in week 52 of 2023 and week 5 of 2024 (Fig. 2B). Among participants testing at least 14 days post-vaccination, 179 (4.6%) tested positive for influenza, of which 153 (85.5%) reported an ARI episode. Among participants who were unvaccinated or under 14 days post-vaccination, 87 (8.1%) tested positive for influenza, of which 74 (85.1%) reported an ARI episode.

### Vaccine effectiveness

The proportional hazards assumption held for all Cox proportional hazards regression models. Model fit was not improved by the addition of covariates, other than those specified a priori. VE was 39.9% (95% CI 21.8 – 53.8) for the primary outcome of influenza infection and 38.7% (18.4 – 53.9) for the secondary outcome of influenza infection with ARI symptoms (Table 2). VE estimates obtained from case-control (41.2%, 22.5 – 55.3) and TNCC (45.9%, 21.8 – 62.2) analyses were similar. Using each approach, 95% CIs around VE estimates overlapped for the periods 14-104 days and ≥105 days post-vaccination (Fig. 3).

**Fig. 3.**
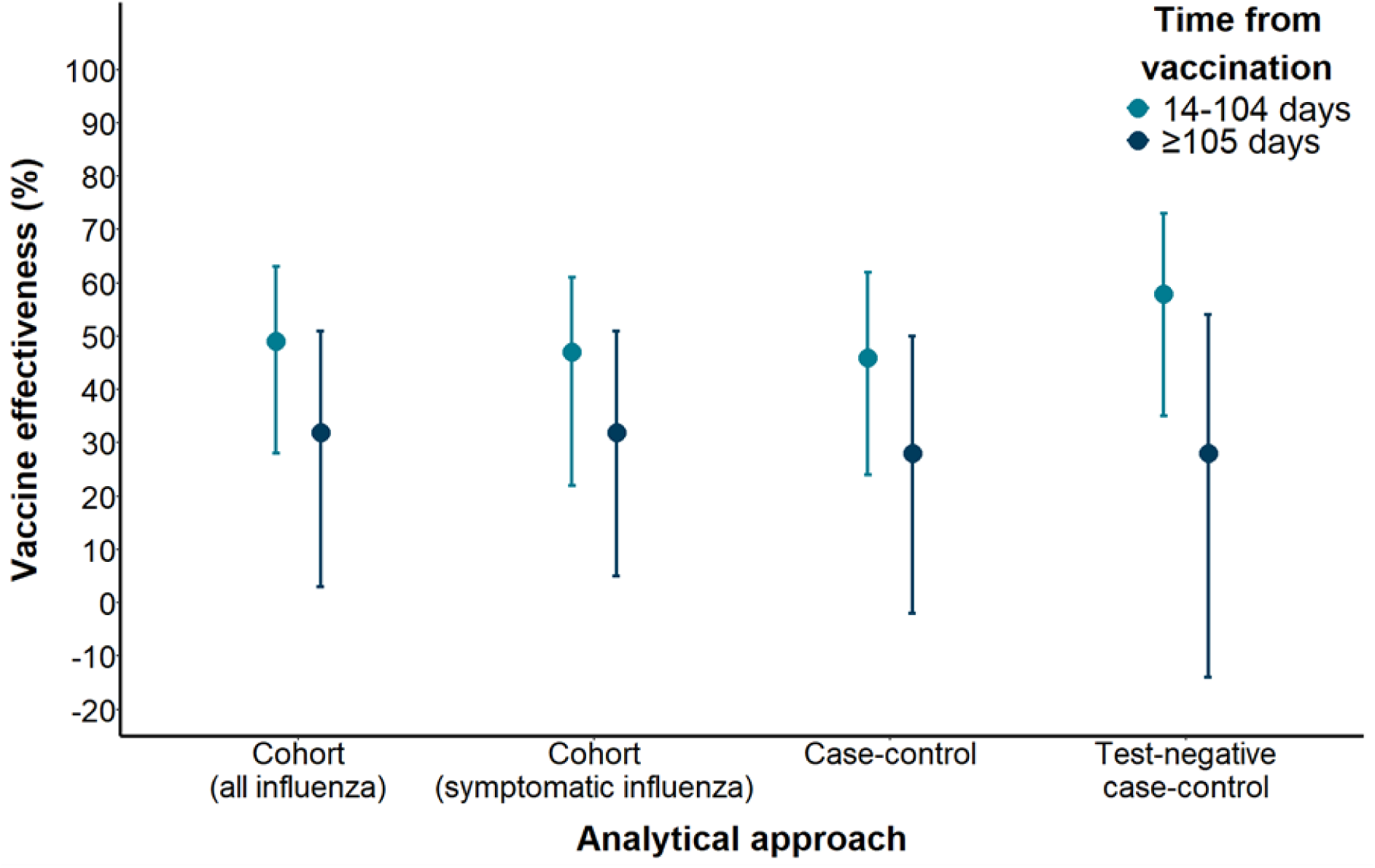
Influenza vaccine effectiveness among SIREN cohort participants during the periods 14-104 days and ≥105 days post-vaccination in the 2023-24 winter season, estimated using cohort, case-control and test-negative case-control analytical approaches. Vaccine effectiveness was estimated using Cox proportional hazards regression for cohort analyses and logistic regression for case-control analyses. Models included age group, sex, presence of chronic disease or immunosuppression, patient-facing role, and region of residence as covariates. Logistic regression models additionally included a spline for number of days since the study period commenced, with 4 degrees of freedom. Symptomatic influenza was defined as an acute respiratory infection (ARI) episode with onset within the period 7 days either side of the sample date of a positive PCR test for any influenza virus, where an ARI episode was defined as a self-report of one or more of cough, sore throat, shortness of breath, or coryza.

**Table 2.** Influenza vaccine effectiveness among SIREN cohort participants during the 2023-24 winter season, estimated using cohort, case-control and test-negative case-control analytical approaches. Vaccine effectiveness was estimated using Cox proportional hazards regression for cohort analyses and logistic regression for case-control analyses. Models included age group, sex, presence of chronic disease or immunosuppression, patient-facing role, and region of residence as covariates. Logistic regression models additionally included a spline for number of days since the study period commenced, with 4 degrees of freedom. * Defined as an acute respiratory infection (ARI) episode with onset within the period 7 days either side of the sample date of a positive PCR test for any influenza virus, where an ARI episode was defined as a self-report of one or more of cough, sore throat, shortness of breath, or coryza. Abbreviations: ARI, acute respiratory infection; PCR, polymerase-chain reaction; VE, vaccine effectiveness.

| Vaccination status (time since vaccination) | Cohort: Time to PCR-positive influenza |  |  | Cohort: Time to PCR-positive influenza with ARI symptoms* |  |  | Case-control |  |  | Test-negative case-control |  |  |
| --- | --- | --- | --- | --- | --- | --- | --- | --- | --- | --- | --- | --- |
|  | Person-days of follow-up | Positive PCR tests | VE (95% CI) | Person-days of follow-up | Positive PCR tests | VE (95% CI) | Cases | Controls | VE (95% CI) | Cases | Controls | VE (95% CI) |
| Unvaccinated | 214,527 | 87 | ref | 188,423 | 73 | ref | 87 | 1,449 | ref | 53 | 1,101 | ref |
| Vaccinated (0-13 days) | 35,161 | 0 | - | 33,262 | 0 | - | 0 | 243 | - | 0 | 162 | - |
| Vaccinated (≥14 days) | 484,291 | 179 | 39.9 (21.8 – 53.8) | 478,479 | 150 | 38.7 (18.4 – 53.9) | 179 | 2,976 | 41.1 (21.9 – 55.3) | 101 | 2,451 | 45.9 (21.8 – 62.2) |
| 14-104 days | 310,150 | 83 | 46.6 (26.5 – 61.2) | 306,546 | 70 | 46.5 (24.4 – 62.1) | 83 | 1,806 | 48.7 (28.2 – 63.3) | 44 | 1,627 | 58.0 (34.9 – 73.0) |
| ≥105 days | 174,141 | 96 | 31.6 (5.3 – 50.6) | 171,933 | 80 | 28.6 (-1.7 – 49.8) | 96 | 1,170 | 31.5 (3.4 – 51.3) | 57 | 824 | 27.6 (-13.7 – 53.7) |

## DISCUSSION

We estimated the effectiveness of seasonal influenza vaccination at preventing laboratory-confirmed infection during the 2023-24 winter season to be 40% in our cohort of UK healthcare workers completing fortnightly PCR testing, 5.4% of whom tested positive for influenza (85% with accompanying ARI symptoms). Similar results were observed across three analytical approaches (cohort, case-control and TNCC).

Applying our results to the wider UK NHS workforce in March 2024, comprising 1.67 million full-time equivalent staff [20–23], it can be crudely estimated that 135,210 staff members would have experienced at least one influenza infection in 2023-24 if they had been unvaccinated, of which 53,950 infections could have been prevented through vaccination. This is important, given the association of HCW infections with workforce absence and nosocomial transmission to patients [2, 3].

### Interpretation of results and comparison with other studies

The UK winter 2023-24 season was characterised by lower and more prolonged influenza activity compared with the 2022-23 season, with co-circulation of the A(H3N2) and A(H1N1)pdm09 subtypes, both belonging to the same clades as vaccine strains [24]. An observed cumulative incidence of influenza in HCWs of 5.4% (4.6% in vaccinated HCWs and 8.1% in unvaccinated HCWs) in this study, lower than previous estimates for HCWs [9–11], may reflect this lower level of influenza activity or the use of different testing methods in previous studies [25]. Peaks in positive influenza PCR tests among participants closely mirrored influenza activity observed nationally [24].

Our VE estimates are similar to results from TNCC studies measuring 2023-24 influenza VE for all laboratory-confirmed influenza among UK adults aged 18-64, including VE against influenza accompanied by: primary care presentation with ARI in Great Britain (49%, 95% 42 – 56), hospitalisation with ARI symptoms in England (31%, 21 – 40), and hospitalisation with a diagnosed respiratory condition in Scotland (48%, 38 – 55) [24]. Our findings are also comparable to those of TNCC studies reporting interim VE estimates against all influenza or influenza A for 2023-24 among adults aged 18-64 years attending healthcare settings in Great Britain, other European countries and North America [26–29]. In each country, influenza vaccination was ordinarily only offered to adults in this age group who had chronic diseases, were pregnant, or had occupational or other specific indications for vaccination. A Cochrane review of the effectiveness of inactivated influenza vaccines among healthy adults, including 25 clinical trials conducted between 1969-2009, reported a pooled VE of 59% (95% CI 53 – 74) [30].

However, a VE of 40% is lower than estimates previously reported among HCWs, though previous studies predominantly used serological testing, which may account in part for these varying estimates. A meta-analysis of two randomised controlled trials and three cohort studies that were conducted between 1992-93 and 2014-15 reported pooled influenza VE for laboratory-confirmed influenza of 64% (46 – 75) among 1,428 HCWs [31]. VE estimates for the individual studies varied between 28% (–55 – 67) and 89% (–105 – 99) [31]. Another meta-analysis included the same three cohort studies and one RCT, reporting a pooled VE for laboratory-confirmed influenza of 60% (31 – 77) overall, 50% (24 – 67) when restricting to observational studies, and 41% (–24 – 72) when restricting to the single study using RT-PCR testing, with evidence of heterogeneity by both study design and modality of testing [32]. Individual studies in these meta-analyses included 72-366 participants per season and most recruited from one or two hospitals. Estimates of influenza incidence and VE would be expected to vary between geographies, influenza seasons, vaccine types and programmes, and study populations and methods.

The design of the SIREN study permitted comparison of VE estimates obtained using different analytical approaches. Our results are in keeping with simulation-based studies reporting consistency in influenza VE estimates obtained using cohort, case-control and TNCC analytical approaches provided that a highly specific diagnostic test is used [13–15].

In this study, 95% CIs for VE during the periods 14-104 days post-vaccination and ≥105 days post-vaccination overlapped, though point estimates were lower for the later period. Existing evidence supports waning VE post-vaccination [33–38], particularly for influenza subtype A/H3 [33, 34, 36], and a decline in antibody titres following seroconversion [33, 39]. Notably, the start date for offering seasonal influenza vaccination of UK HCWs and selected other population groups was moved to 1 October rather than 1 September for the 2024-25 influenza season onwards in light of waning of VE [40].

### Strengths and weaknesses of study

A key strength of this study was the availability of data from serial PCR tests and questionnaires, with very good participant adherence to the study protocol, enabling analysis of VE against all influenza infection, not just symptomatic cases. Additionally, we were able to assess the consistency of VE estimates across different analytical approaches. Given routine completion of respiratory samples and symptom questionnaires irrespective of symptom status, our findings are unlikely to be affected by bias by healthcare-seeking behaviour.

This study also had several limitations. First, the 14-day interval between successive respiratory swabs for PCR, selected based on testing for SARS-CoV-2, means that some participants could have developed and cleared influenza infections, particularly shorter and less severe infections, between successive swabs. This could have led to underascertainment of influenza infections.

Second, data was not available on positivity for influenza types and subtypes, and we had limited power to conduct subgroup analyses, resulting in uncertainty around VE estimates for shorter post-vaccination periods and an inability to assess VE by vaccine type.

Third, vaccine uptake among study participants exceeded that among UK HCWs in general, though this would not necessarily be expected to affect the generalisability of VE estimates.

## CONCLUSION

The findings of this study, the largest to our knowledge to estimate influenza VE in HCWs, emphasise the ongoing importance of routine seasonal influenza vaccination of this occupational group. When scaled to the level of the overall UK healthcare workforce, an observed VE of 40% in 2023-24 could prevent a substantial burden of infection in HCWs, as well as reducing impacts on patients and workforce pressures.

Evidence suggests that low uptake of seasonal influenza vaccination among healthcare staff is in part related to perceptions regarding a lack of personal infection risk and poor vaccine efficacy [8].

Consequently, communication within vaccination campaigns of information directly relevant to HCWs, together with the potential benefits for patients and the wider healthcare system, may help to support vaccine uptake. However, efforts to increase influenza vaccine uptake in HCWs are likely to require a multifaceted approach that includes a focus on supply of and access to vaccines, as well as educational approaches to address existing concerns.

## Data Availability

The anonymised SIREN dataset will be made available for secondary analysis to approved researchers upon reasonable request, subject to meeting appropriate information governance requirements.

## ACKNOWLEDGEMENTS

We thank all the participants for their ongoing contributions and commitment to this study, and the research teams at all 89 SIREN sites actively following up participants during this analysis period for their support and for making the study possible. We thank Diane Corrigan at Public Health Agency Northern Ireland, Kevin Wilson at Public Health Scotland, Elen de Lacy at Public Health Wales, and Chris Norman at Health and Care Research Wales for supporting SIREN delivery and providing data from their national immunisation registries.

